# Cognitive reserve and gamma entrainment-related network changes in prodromal Alzheimer’s disease

**DOI:** 10.64898/2026.04.28.26351913

**Authors:** Myeonghae Hyun, Yeseung Park, Euisuk Yoon, Yejung Kim, Hyeonwook Chae, Seunghyup Yoo, Ji Won Han, Ki Woong Kim

## Abstract

**Background and Objectives:** Gamma Entrainment responses in Alzheimer’s disease are heterogeneous, but factors underlying this variability remain unclear. Cognitive reserve may influence how large-scale brain networks reconfigure during stimulation, particularly in prodromal Alzheimer’s disease. We examined whether cognitive reserve was associated with stimulation-related functional network changes across the Alzheimer’s disease spectrum and hypothesized a condition-by-diagnosis-by-cognitive reserve interaction.

**Methods:** In this cross-sectional EEG study, adults aged 55 years or older were recruited from a memory clinic and community. Participants underwent amyloid PET and standardized neuropsychiatric and neuropsychological assessment and were classified as amyloid-negative cognitively normal, amyloid-positive mild cognitive impairment, or amyloid-positive Alzheimer’s disease dementia. Amyloid-negative mild cognitive impairment and amyloid-positive cognitively normal participants were excluded from the primary analyses. Cognitive reserve was indexed using an education- and occupation-based composite and dichotomized at the median. Functional networks during rest and 32-Hz visual stimulation were derived from spectral Granger causality EEG and summarized using mean strength, weighted clustering coefficient, average shortest path length, and outreach. Mixed-design repeated-measure ANCOVA adjusted for age, sex, and APOE genotype was used for the primary analysis.

**Results:** Significant condition-by-diagnosis-by-cognitive reserve interactions were observed for mean strength, weighted clustering coefficient, and outreach, but not for average shortest path length. These effects were driven primarily by prodromal Alzheimer’s disease, in which the low cognitive reserve subgroup showed larger rest-to-stimulation increases than the high cognitive reserve subgroup for mean strength, weighted clustering coefficient, and outreach. In secondary analyses, the low cognitive reserve prodromal subgroup also showed higher 32-Hz entrainment, and entrainment strength correlated positively with stimulation-related network change across the full sample.

**Discussion:** Cognitive reserve may be associated with heterogeneity in stimulation-related network reconfiguration, with the clearest signal observed in prodromal Alzheimer’s disease. Because the low-reserve prodromal subgroup was small and findings were sensitive to alternative modeling choices, these results should be considered preliminary. Reserve-related factors may warrant explicit consideration in future studies of sensory gamma stimulation in Alzheimer’s disease.

## Introduction

In recent years, rhythmic sensory stimulation has gained attention as a potential neuromodulatory approach for Alzheimer’s disease (AD), particularly in relation to gamma-frequency activity^1–3^. Gamma oscillations, typically spanning the 30-80 Hz range, are involved in higher-order cognitive processes such as attention^4^, memory^5^, and perception^6^, and growing evidence suggests that these rhythms are disrupted in AD^7, 8^. This has led to increasing interest in gamma entrainment, defined as the synchronization of ongoing neural activity to an external rhythmic stimulus delivered at gamma frequencies^9–11^. Preclinical studies have further supported the therapeutic potential of gamma entrainment by showing that sensory stimulation at gamma frequencies, most notably 40 Hz flicker paradigms, can reduce AD-related pathology and improve behavioral outcomes in transgenic mouse models^11–13^. However, translational findings from human studies have been more heterogeneous, suggesting that individual factors may substantially influence the neural response to sensory gamma stimulation^10, 14^.

AD is increasingly understood not only as a disorder of regional pathology but also as a network-level disorder characterized by disrupted communication across large-scale brain systems^15, 16^. Amyloid-β and tau pathology progressively impair synaptic integrity and weaken interregional coordination, contributing to altered patterns of functional integration and segregation^17, 18^. From this perspective, evaluating the effects of gamma stimulation requires more than confirming local entrainment alone; it also requires assessing how stimulation reshapes network organization at the systems level. Graph theory provides a useful framework for this purpose by quantifying network properties related to integration, segregation, and communication efficiency across the whole brain^19–21^. In addition, directed connectivity measures offer an important advantage because they preserve information about the direction of information flow between brain regions. Spectral Granger causality (sGC), a frequency-domain measure of directed influence, is therefore well suited for capturing stimulation-related changes in oscillatory EEG networks^22, 23^. In the present study, we combined sGC with graph-theoretical analysis to characterize stimulation-related changes in brain network topology, including mean strength, weighted clustering coefficient, average shortest path length, and outreach. Although sGC captures directed causal influences, the primary graph analyses were based on symmetrized connectivity matrices to facilitate standard graph-theoretical characterization; a confirmatory directed reanalysis was performed separately to verify that this step did not alter the findings.

Cognitive reserve (CR) is a key construct for understanding why individuals with similar levels of neuropathology may show different degrees of cognitive impairment^24–26^. Rather than reflecting the absence of pathology, CR refers to the capacity to cope with brain changes through more efficient, flexible, and compensatory use of neural resources^24–26^. Experiences such as education, occupational attainment, and cognitively enriching activities are thought to contribute to this capacity and have been associated with greater neural efficiency, adaptive network recruitment, and resilience to clinical decline^24, 27^. If CR reflects differences in the efficiency and flexibility of large-scale network recruitment, it may also help explain why individuals with similar levels of AD pathology show heterogeneous neural responses to external gamma-frequency stimulation. This question may be especially relevant in prodromal Alzheimer’s disease (PAD), defined here as amyloid-positive mild cognitive impairment, because PAD represents a transitional stage in which AD pathology is already present but compensatory network mechanisms may still be sufficiently active to shape the brain response to stimulation. By contrast, reserve-related differences may be less apparent in cognitively normal individuals with lower pathological burden or in later-stage AD, where network disruption may be too advanced to support flexible reconfiguration. In the present study, CR was indexed using a composite measure based on education and occupational attainment, following the approach described by Garibotto and colleagues^28, 29^.

To our knowledge, no previous study has directly tested whether cognitive reserve is associated with differences in stimulation-related functional brain network reconfiguration during visual gamma entrainment across the AD spectrum. In the present study, we applied 32 Hz visual flicker stimulation and evaluated stimulation-related changes in sGC-derived functional network topology in cognitively normal individuals, patients with PAD, and patients with clinical AD. We hypothesized that stimulation-related changes in functional network topology would differ according to both diagnostic stage and cognitive reserve, with the greatest heterogeneity expected in the prodromal stage of AD. Specifically, we predicted a three-way interaction among condition, diagnostic status, and cognitive reserve. Finally, as an exploratory analysis, we examined whether PAD participants with low and high cognitive reserve would show distinct patterns of network reconfiguration in response to stimulation.

## Materials and methods

### Participants

We recruited adults aged 55 years or older from the memory clinic at Seoul National University Bundang Hospital and from community-dwelling older adults between April 2023 and June 2024. This study was approved by the Institutional Review Board of Seoul National University Bundang Hospital (IRB No. B-2304-823-001) and was conducted in accordance with the Declaration of Helsinki. All participants provided written informed consent prior to enrolment.

Inclusion criteria were as follows: age 55 years or older, completion of amyloid PET imaging, and availability of standardized clinical neuropsychological evaluation sufficient for diagnostic classification. Based on amyloid PET status and clinical diagnosis, participants were categorized as amyloid-negative cognitively normal (CN), amyloid-positive mild cognitive impairment (prodromal AD, PAD), or amyloid-positive dementia due to Alzheimer’s disease (AD). Exclusion criteria included amyloid-negative mild cognitive impairment (*n*=13), because cognitive symptoms in this group may reflect non-Alzheimer etiologies, and amyloid-positive cognitively normal (*n*=12) status in the primary analyses, because the available sample size was insufficient for stable estimation in diagnosis-by-cognitive reserve subgroup models. An initial pool of 120 participants was enrolled (32 men, 88 women; mean age, 74.3 ± 6.9 years). The final analytic sample comprised 95 participants (22 men, 73 women; mean age 74.4 ± 6.8 years), including amyloid-negative cognitively normal controls and the symptomatic AD spectrum (PAD and AD).

### Clinical Assessment and Cognitive Reserve Assessment

All participants underwent standardized clinical evaluations by geriatric psychiatrists, including medical and neurological examinations, laboratory testing, and neuropsychological assessment using the Korean version of the Consortium to Establish a Registry for Alzheimer’s Disease Assessment Packet (CERAD-K)^30^, along with psychiatric screening using the Korean version of the Mini International Neuropsychiatric Interview (K-MINI)^31^. Diagnostic classification was established by dementia specialists based on amyloid PET findings and standardized clinical and neuropsychological evaluations. Cognitively normal participants were defined as amyloid-negative individuals who performed no lower than 1.5 standard deviations below age-, sex-, and education-adjusted norms on all CERAD-K tests and maintained independence in basic and instrumental activities of daily living. Mild cognitive impairment was diagnosed according to the International Working Group (IWG) criteria for mild cognitive impairment, defined as objective cognitive impairment with performance below 1.5 standard deviations of age-, sex-, and education-adjusted norms on neuropsychological testing while preserving independence in daily functioning^32^. Prodromal AD was defined as amyloid-positive mild cognitive impairment. Dementia was diagnosed according to the DSM-IV, and dementia due to Alzheimer’s disease additionally met the National Institute on Aging-Alzheimer’s Association (NIA-AA) criteria for probable Alzheimer’s disease^33^. The AD group comprised amyloid-positive participants who fulfilled these criteria.

Cognitive reserve (CR) was quantified using an education- and occupation-based composite proxy score, following the reserve-proxy approach described by Garibotto *et al*. ^28, 29^. Education was defined as completed years of formal schooling and converted to a 6-level ordinal score based on the distribution of education years in our sample, thereby placing it on the same scale as the occupational proxy. Education categories were scored as follows: 1, less than elementary school; 2, elementary school graduate to before middle school graduation; 3, middle school graduate to before high school graduation; 4, high school graduate; 5, junior college to university graduate; and 6, postgraduate education. Occupational attainment was coded on a 6-point scale according to the participants’ last occupation as follows: 1, no occupation; 2, unskilled labourer; 3, housewife; 4, skilled labor/tradesman/lower-level civil servant or employee/self-employed with a small business/office or sales work; 5, mid-level civil servant or management/head of a small business/academic or specialist in a subordinate position; and 6, senior civil servant or senior management/senior academic position/self-employed with high responsibility. The final CR score was calculated as the sum of the education score (1-6) and occupation score (1-6). Participants were dichotomized into low-CR (LCR) and high-CR (HCR) groups using a median split of the CR score in the analytic sample. This education- and occupation-based proxy was chosen for its established validity in studies of cognitive reserve across the Alzheimer’s disease spectrum^28, 29^. More comprehensive instruments incorporating leisure and social activities were not available in the present cohort.

### Visual flicker stimulation protocol

We developed a custom eyewear device (A88MA2B; Konica Minolta Inc., Tokyo, Japan) delivering square-wave modulated white light (5700K color temperature, 1120 cd/m² luminance, 50% duty cycle) at programmable frequencies (32, 34, 36, 38, and 40 Hz), positioned 2 cm from each pupil. Device safety features are described in **Supplementary Appendix 1**.

For EEG recording sessions, the device operation was modified to enable precise experimental control. A function generator (TG 5012A, Aim & Thurlby Thandar Instruments, Huntingdon, UK) replaced the standard AC circuit to ensure exact timing for steady-state visual evoked potential (SSVEP) measurements. Each EEG session began with a 5-minute resting-state recording, followed by four stimulation blocks separated by 30-second breaks. Within each stimulation block (370 seconds), five frequency sub-blocks (32, 34, 36, 38, and 40 Hz) were presented in a randomized order, separated by 10-second intervals. Each frequency sub-block consisted of five trials. Individual trials lasted 8 seconds with randomized inter-trial intervals of 6.5 ± 1.5 seconds to minimize habituation effects.

### EEG acquisition, preprocessing, and network analysis

EEG data were recorded using 64-channel Ag–AgCl electrode caps (Easycap, EASYCAP GmbH, Munich, Germany) according to the extended International 10–20 System, with FCz as reference, at a sampling rate of 1,000 Hz using a 24-bit ActiCHamp DC amplifier and recorded with BrainVision Recorder software (Brain Products GmbH, Gilching, Germany). Preprocessing was performed in MATLAB using EEGLAB and BSMART toolboxes. Signals were filtered (1-Hz high-pass, 60-Hz notch), re-referenced to common average, and artefact-corrected using independent component analysis with conservative rejection (capped at 20% of components per participant).

Pairwise sGC was computed for all unique channel pairs among 63 EEG channels (1,953 pairs) using BSMART toolbox. sGC was estimated with a fixed model order of 75, a sampling rate of 1,000 Hz, and an analysis window of 7,500 samples (7.5 s). The model order of 75, corresponding to 75 ms, was chosen to capture approximately 2-3 cycles of gamma-band activity in the 32-40 Hz range^34^. For resting-state analysis, the 5-minute recordings were divided into 1,500-ms epochs, from which 20 artefact-free epochs were randomly selected. For stimulation trials, although each visual stimulation trial lasted 8 s, EEG data were epoched into 10-s segments spanning 1,000 ms before stimulus onset to 1,000 ms after stimulus offset. To minimize onset-related transients while capturing sustained steady-state responses, sGC was computed from a 7.5-s window corresponding to 501-8,000 ms after stimulus onset (samples 1,501-9,000 within each 10-s epoch). Spectral estimates were computed from 30 to 42 Hz in 1-Hz steps, and frequency-resolved sGC values were assembled into a 63 X 63 X 13 matrix. Although sGC was computed for all five stimulation frequencies, primary analyses were based on the 32 Hz condition. The original asymmetric sGC matrices were preserved for secondary directed analyses. Additionally, 32 Hz SNR values were extracted to assess entrainment strength, and resting-state relative power was calculated across six frequency bands for baseline spectral comparisons.

Functional brain networks were constructed from the sGC matrices, with 63 EEG channels as network nodes and sGC values as weighted edges. Self-connections were set to zero and sGC matrices were symmetrized by averaging reciprocal connections; a directed reanalysis of the raw asymmetric matrices was performed separately to verify this step **(Supplementary Table S8)**. A proportional thresholding strategy was applied, with densities explored from 45% to 100% in 5% increments in the full enrolled dataset (*N* = 120). A density of 90% was selected for the primary analyses to retain the majority of connectivity information while removing the weaker connections most likely to reflect noise. This threshold was determined prior to and independently of the statistical comparisons of interest; no group-level effects were examined during threshold selection. Because sGC captures directional frequency-specific coupling and produces non-zero values for a large proportion of channel pairs, the resulting connectivity matrices are inherently denser than those obtained from correlation-based approaches, and conventional lower thresholds (e.g. 10-30%) would discard a substantial proportion of physiologically meaningful connections. The selected threshold was applied unchanged to the final analytic sample (*N* = 95). Sensitivity analyses at 50% and 70% densities were also conducted.

Graph theoretical metrics were calculated for the thresholded networks using the Brain Connectivity Toolbox. We computed four key measures. Mean Strength (MS) was defined as the average node strength (sum of edge weights per node) and was averaged across nodes to obtain a global summary. Weighted Clustering Coefficient (Cw) quantified weighted functional segregation. Characteristic Path Length (PL) was computed after converting edge weights to connection lengths using a standard weight-to-length transformation, followed by computation of weighted shortest paths and their average across node pairs. Outreach (OUT) was defined as the node-wise sum of edge weights multiplied by the Euclidean physical distance between electrode pairs (OUT□ = Σ□ w□□ × d□□) and was averaged across nodes to obtain a global outreach estimate. Outreach was included to complement purely topological metrics with a spatially informed measure, as cognitive reserve effects on stimulation-related network changes may manifest not only in connection strength and clustering but also in the spatial extent of network engagement.

All analyses were performed using MATLAB with EEGLAB, BSMART and BCT toolboxes.

### Statistical Analysis

Participants’ demographic and clinical characteristics were compared across diagnostic groups using one-way analysis of variance for continuous variables and Pearson’s chi-square test for categorical variables.

The primary network analysis used a single prespecified 32 Hz rest-versus-stimulation contrast, grounded in our prior parameter-optimization work showing that 32 Hz most effectively entrained gamma rhythms in older adults^35–37^. Mixed-design repeated-measures ANCOVA models were fitted with Condition (rest, stimulation) as a within-subject factor, diagnostic stage (CN, PAD, AD) and cognitive reserve (LCR, HCR) as between-subject factors, and age, sex, and APOE genotype as covariates. An a *priori* power analysis (G*Power 3.1; *f* = 0.25, α= 0.05, power = 0.80) estimated a minimum of 60 participants; the final sample of 95 exceeded this requirement.

Benjamini-Hochberg FDR correction was applied across the four three-way interaction tests. Two-way interactions and main effects were evaluated as part of the same omnibus ANCOVA model and are reported at nominal significance levels, as these were interpreted only in conjunction with significant three-way interactions. Follow-up within-group ANCOVAs were conducted only for metrics that survived FDR correction at the omnibus level, and post-hoc pairwise comparisons within these models were Bonferroni-adjusted. Model-based standardized effect sizes (d_adjusted = ΔEMM/√MSE_RS) were computed for each rest-versus-stimulation contrast; because these are derived from covariate-adjusted residual variance rather than raw pooled standard deviations, they may appear larger than conventional Cohen’s *d*, particularly in small subgroups. Unadjusted effect sizes are provided in **Supplementary Appendix 2**.

Supportive analyses included: (i) Welch’s t-tests comparing 32 Hz SNR between CR subgroups within each diagnostic stage; (ii) Pearson correlations between 32 Hz SNR and stimulation-related Δ sGC; (iii) resting-state relative power comparisons between PAD CR subgroups; (iv) threshold sensitivity at 50%, 70%, and 90% densities; (v) permutation testing with 10,000 permutations; and (vi) directed-network reanalysis from the raw asymmetric sGC matrices. Exploratory analyses examined the ΔPL/ΔMS ratio, OUT/MS ratio, and quadrant-based response-pattern classification within PAD; *p* values for these analyses are reported without multiplicity correction given their hypothesis-generating nature. A total of three exploratory tests were conducted (ΔPL/ΔMS ratio, OUT/MS ratio, and quadrant-based classification); no additional exploratory tests were performed and subsequently omitted.

## Results

A total of 95 participants were included in the present study and were categorized into CN, PAD, and AD groups. Baseline comparisons showed that age, Mini-Mental State Examination (MMSE) score, cognitive reserve score, and APOE ε4 carrier status differed across diagnostic groups **(Table 1)**, reflecting clinical differences between groups. Accordingly, all subsequent analyses controlled for age, sex, and APOE ε4 status.

**Table 1.**
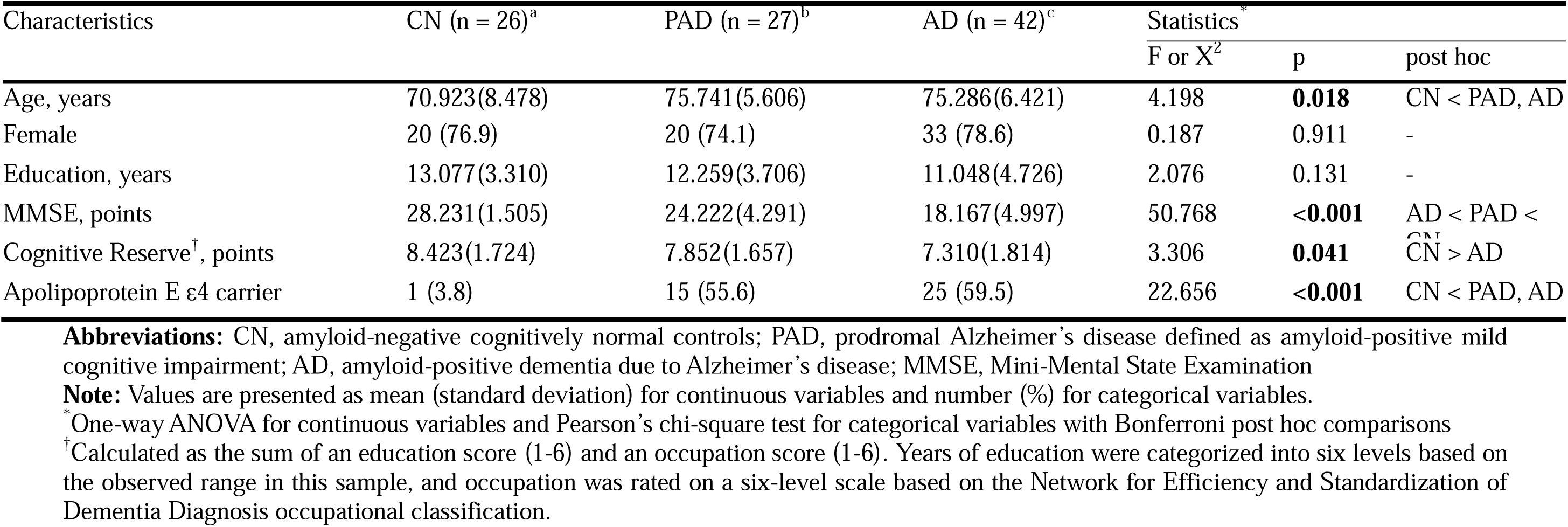
Baseline characteristics of participants.

Cognitive reserve was operationalized using education rank and occupation rank, which were summed to yield a total cognitive reserve score. Education rank ranged from 1 to 6 (mean 3.89, SD 1.28, median 4) and was most frequent at ranks 4 (*n* = 28, 29.5%) and 5 (*n* = 30, 31.6%). Occupation rank ranged from 2 to 6 (mean 3.87, SD 0.88, median 4) and was most frequent at rank 4 (*n* = 45, 47.4%). The total score ranged from 3 to 12 (mean 7.77, SD 1.79, median 8), with most participants scoring between 6 and 9 (*n* = 71, 74.4%). The distribution of cognitive reserve scores across diagnostic groups and the median split threshold are illustrated in **Supplementary Figure S5**.

At baseline during the rest condition, graph-theoretical network metrics were largely comparable across diagnostic and cognitive reserve groups. As shown in **Table 2** and **Supplementary Table S1**, no significant main effects of diagnosis or cognitive reserve were observed for most metrics, including mean strength, weighted clustering coefficient, and outreach, and there was no evidence of a diagnosis by cognitive reserve interaction at rest. Although average shortest path length showed some variation across diagnostic groups, this effect did not reach statistical significance. Overall, these findings suggest that group differences were less apparent at rest and became more observable during stimulation-related network responses.

**Table 2.** Graph metrics of functional connectivity at rest and during stimulation by diagnosis and cognitive reserve.

|  | CN (n = 26) |  | PAD (n = 27) |  | AD (n = 42) |  |
| --- | --- | --- | --- | --- | --- | --- |
|  | LCR(n = 6) | HCR(n = 20) | LCR(n = 9) | HCR(n = 18) | LCR(n = 23) | HCR(n = 19) |
| Mean strength |  |  |  |  |  |  |
| Rest | 2.315(1.273) | 2.512(1.699) | 3.052(1.133) | 2.419(1.219) | 2.492(1.921) | 2.132(0.876) |
| Stimulation | 5.623(2.092) | 5.943(2.823) | 11.304(6.943) | 5.653(4.394) | 5.092(2.870) | 4.984(3.234) |
| Weighted clustering coefficient |  |  |  |  |  |  |
| Rest | 0.031(0.018) | 0.032(0.018) | 0.040(0.015) | 0.032(0.017) | 0.034(0.028) | 0.029(0.012) |
| Stimulation | 0.080(0.030) | 0.081(0.038) | 0.153(0.100) | 0.077(0.064) | 0.071(0.042) | 0.068(0.046) |
| Average shortest path length |  |  |  |  |  |  |
| Rest | 25.009(8.506) | 27.737(10.846) | 19.728(8.072) | 28.980(15.603) | 32.887(16.124) | 29.750(10.294) |
| Stimulation | 14.314(13.075) | 11.443(6.194) | 6.290(3.086) | 15.913(10.122) | 14.424(7.224) | 16.279(9.919) |
| Outreach |  |  |  |  |  |  |
| Rest | 1.300(0.753) | 1.335(1.078) | 1.702(0.628) | 1.290(0.660) | 1.323(1.014) | 1.171(0.537) |
| Stimulation | 3.132(1.215) | 3.302(1.655) | 6.577(4.016) | 3.175(2.573) | 2.771(1.576) | 2.774(1.916) |

**Abbreviations:** CN, amyloid-negative cognitively normal controls; PAD, prodromal Alzheimer's disease defined as amyloid-positive mild cognitive impairment; AD, amyloid-positive dementia due to Alzheimer's disease; LCR, low cognitive reserve; HCR, high cognitive reserve
**Note:** Values are presented as mean (standard deviation).

We next conducted a mixed-design repeated-measures ANCOVA to examine the effects of condition (rest vs. stimulation), diagnosis, and cognitive reserve on each network metric **(Table 3)**. A significant main effect of condition was observed for weighted clustering coefficient (*F*_(1,86)_ = 4.574, *p* = 0.035), whereas the corresponding condition effects for mean strength (*F*_(1,86)_ = 3.855, *p* = 0.053) and outreach (*F*_(1,86)_ = 3.299, *p* = 0.073) did not reach statistical significance. Diagnosis showed robust main effects on mean strength (*F*_(2,86)_ = 6.780, *p* = 0.002), weighted clustering coefficient (*F*_(2,86)_ = 5.758, *p* = 0.005), average shortest path length (*F*_(2,86)_ = 3.554, *p* = 0.033), and outreach (*F*_(2,86)_ = 7.652, *p* = 0.001). In contrast, the main effect of cognitive reserve was not statistically significant for any of the examined metrics, including mean strength (*F*_(1,86)_ = 2.916, *p* = 0.091), weighted clustering coefficient (*F*_(1,86)_ = 3.214, *p* = 0.077), average shortest path length (*F*_(1,86)_ = 0.446, *p* = 0.506), and outreach (*F*_(1,86)_ = 3.144, *p* = 0.080).

**Table 3.** Effects of condition, diagnosis, and cognitive reserve on the graph metrics of functional connectivity*.

|  | MS |  |  | Cw |  |  | PL |  |  | Outreach |  |  |
| --- | --- | --- | --- | --- | --- | --- | --- | --- | --- | --- | --- | --- |
| | F | p | $\eta p^2$ | F | p | $\eta p^2$ | F | p | $\eta p^2$ | F | p | $\eta p^2$ |
| Condition | 3.855 | 0.053 | 0.043 | 4.574 | **0.035** | 0.050 | 1.525 | 0.220 | 0.017 | 3.299 | 0.073 | 0.037 |
| Diagnosis | 6.780 | **0.002** | 0.136 | 5.758 | **0.005** | 0.118 | 3.554 | **0.033** | 0.076 | 7.652 | **0.001** | 0.151 |
| Cognitive reserve | 2.916 | 0.091 | 0.033 | 3.214 | 0.077 | 0.036 | 0.446 | 0.506 | 0.005 | 3.144 | 0.080 | 0.035 |
| Condition*Diagnosis | 5.515 | **0.006** | 0.114 | 5.093 | **0.008** | 0.106 | 0.348 | 0.707 | 0.008 | 6.125 | **0.003** | 0.125 |
| Condition*Cognitive reserve | 3.673 | 0.059 | 0.041 | 3.793 | 0.055 | 0.042 | 0.543 | 0.463 | 0.006 | 3.536 | 0.063 | 0.039 |
| Diagnosis*Cognitive reserve | 4.009 | **0.022** | 0.085 | 3.216 | **0.045** | 0.070 | 2.156 | 0.122 | 0.048 | 4.631 | **0.012** | 0.097 |
| Condition*Diagnosis*Cognitive reserve | 4.624 | **0.012** | 0.097 | 4.183 | **0.018** | 0.089 | 1.306 | 0.276 | 0.029 | 4.828 | **0.010** | 0.101 |

**Abbreviations:** MS, mean strength; Cw, weighted clustering coefficient; PL, average shortest path length
\*Analysis of variance examined the main effects of condition (resting versus stimulation), diagnosis (amyloid-negative cognitively normal controls versus prodromal Alzheimer's disease versus amyloid-positive dementia due to Alzheimer's disease), and cognitive reserve (low versus high), and their interactions on each graph metric

Two-way interaction analyses were performed to determine whether stimulation-related network changes differed by diagnostic stage and cognitive reserve. The interaction between condition and diagnosis was significant for mean strength (*F*_(2,86)_ = 5.515, *p* = 0.006), weighted clustering coefficient (*F*_(2,86)_ = 5.093, *p* = 0.008), and outreach (*F*_(2,86)_ = 6.125, *p* = 0.003). The interaction between diagnosis and cognitive reserve was also significant for mean strength (*F*_(2,86)_ = 4.009, *p* = 0.022), weighted clustering coefficient (*F*_(2,86)_ = 3.216, *p* = 0.045), and outreach (*F*_(2,86)_ = 4.631, *p* = 0.012). The interaction between condition and cognitive reserve did not reach statistical significance for any of the examined metrics, including mean strength (*F*_(1,86)_ = 3.673, *p* = 0.059), weighted clustering coefficient (*F*_(1,86)_ = 3.793, *p* = 0.055), average shortest path length (*F*_(1,86)_ = 0.543, *p* = 0.463), and outreach (*F*_(1,86)_ = 3.536, *p* = 0.063).

Most importantly, significant three-way interactions among condition, diagnosis, and cognitive reserve were observed for mean strength (*F*_(2,86)_ = 4.624, *p* = 0.012), weighted clustering coefficient (*F*_(2,86)_ = 4.183, *p* = 0.018), and outreach (*F*_(2,86)_ = 4.828, *p* = 0.010). This indicates that the relationship between cognitive reserve and stimulation-related network change depended on diagnostic stage. In contrast, the three-way interaction was not significant for average shortest path length (*F*(2,86) = 1.306, *p* = 0.276), suggesting that the condition by cognitive reserve pattern observed for the other metrics did not extend to this measure of functional integration. **Fig. 1** illustrates these condition-specific response patterns across diagnostic and cognitive reserve groups. This pattern remained unchanged after false discovery rate correction, with significant three-way interactions for mean strength, weighted clustering coefficient, and outreach (all adjusted *p* = 0.024), but not for average shortest path length (adjusted *p* = 0.276).

**Figure 1.**
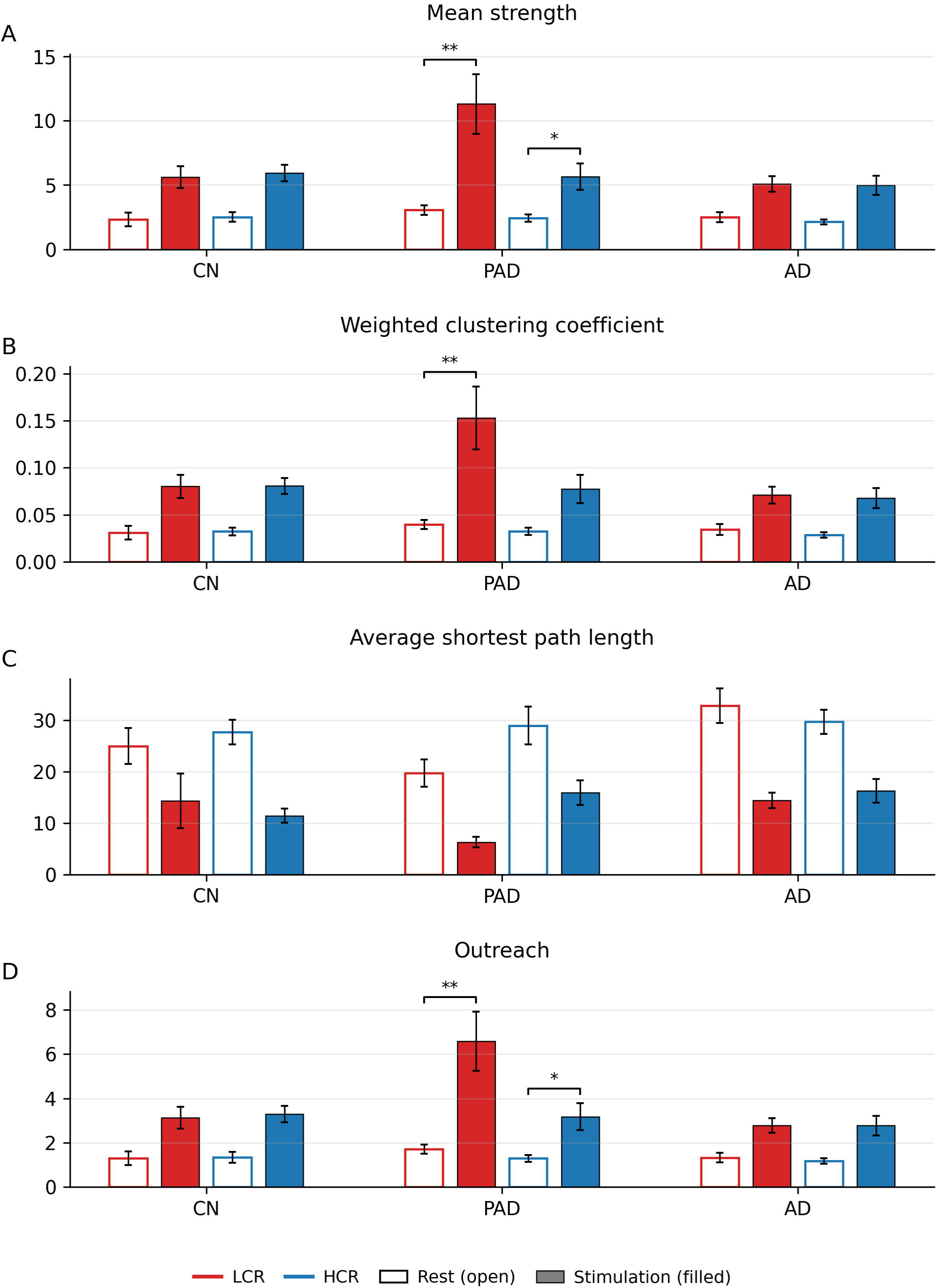
Graph-theoretical metrics at rest and during stimulation across diagnostic groups and cognitive reserve groups. Mean strength, weighted clustering coefficient, average shortest path length, and outreach are shown for CN, PAD, and AD groups, stratified by cognitive reserve. Bars represent covariate-adjusted group means and error bars indicate standard error. Significance brackets with asterisks indicate covariate-adjusted post hoc comparisons between stimulation and rest within each cognitive reserve subgroup separately in each diagnostic group. **Abbreviations:** CN, amyloid-negative cognitively normal controls; PAD, prodromal Alzheimer’s disease defined as amyloid-positive mild cognitive impairment; AD, amyloid-positive dementia due to Alzheimer’s disease; LCR, low cognitive reserve; HCR, high cognitive reserve. *p<.05, **p<.001.

As a supplementary frequency-specific check, the same model was repeated at 34 Hz; the corresponding three-way interactions were weaker overall and are reported in **Supplementary Table S4**.

To identify which diagnostic group drove these three-way interactions, follow-up two-way repeated-measures ANCOVAs were conducted separately within each diagnostic group. Notably, the PAD group was the only diagnostic group to show a significant condition-by-cognitive reserve interaction for the three metrics that showed significant omnibus three-way interactions (**Supplementary Table S2**), namely mean strength (*F*_(1,22)_ = 15.565, *p* = 0.001), weighted clustering coefficient (*F*_(1,22)_ = 15.561, *p* = 0.001), and outreach (*F*_(1,22)_ = 13.864, *p* = 0.001). In contrast, neither the CN nor the AD group showed a significant condition-by-cognitive reserve interaction, indicating a comparable CR-related stimulation pattern was not detected in those groups.

Post hoc simple-effect analyses within the PAD group further clarified this divergence. LCR subgroup showed robust increases from rest to stimulation in mean strength (*t*(22) = 6.60, adjusted *p* < 0.001, d_adjusted = 3.69, 95% CI [2.72, 6.51]; unadjusted *d* = 1.30), weighted clustering coefficient (*t*(22) = 6.59, adjusted *p* < 0.001, d_adjusted = 3.64, 95% CI [2.60, 6.65]; unadjusted *d* = 1.23), and outreach (*t*(22) = 6.34, adjusted *p* < 0.001, d_adjusted = 3.55, 95% CI [2.57, 6.21]; unadjusted *d* = 1.33), indicating larger rest-to-stimulation increases in these network metrics within the PAD-LCR subgroup. The HCR subgroup also showed smaller but significant increases in mean strength (*t*(22) = 2.18, adjusted *p* = 0.040, d_adjusted = 0.80, 95% CI [0.16, 1.77]; unadjusted *d* = 0.74) and outreach (*t*(22) = 2.24, adjusted *p* = 0.036, d_adjusted = 0.82, 95% CI [0.17, 1.81]; unadjusted *d* = 0.74), whereas the change in weighted clustering coefficient did not reach significance (*t*(22) = 1.93, adjusted *p* = 0.060, d_adjusted = 0.73, 95% CI [0.10, 1.71]; unadjusted *d* = 0.70). Collectively, these results suggest that stimulation-related network amplification within PAD appeared greater in the LCR subgroup than in the HCR subgroup. Unadjusted within-subject effect sizes confirmed a consistent LCR > HCR pattern (LCR: *d* = 1.30, 1.23, 1.33; HCR: *d* = 0.74, 0.70, 0.74 for mean strength, weighted clustering coefficient, and outreach, respectively; Supplementary Appendix 2). Because the PAD-LCR subgroup was small, the large adjusted effect sizes, particularly for mean strength, should be interpreted with caution given the small subgroup size. Pairwise p values were Bonferroni-adjusted, and 95% CIs for d_adjusted were estimated using stratified bootstrap resampling with 5000 iterations.

The following supportive analyses were prespecified to evaluate the physiological plausibility and analytic robustness of the primary findings. The PAD-LCR subgroup had higher 32 Hz SNR than the PAD-HCR subgroup (mean = 2.729 vs. 1.230, *p* = 0.026, *d* = 1.015), whereas no cognitive reserve-related SNR differences were observed in the CN or AD groups **(Supplementary Table S7)**. Moreover, across the full sample, 32 Hz SNR was positively correlated with the magnitude of stimulation-related network changes (*r* = 0.360, *p* < 0.001), and this association was also observed within the CN group (*r* = 0.522, *p* = 0.006) and the AD group (*r* = 0.307, *p* = 0.048), with a trend in the same direction within PAD (*r* = 0.353, *p* = 0.071) **(Supplementary Figure S4)**. This entrainment-network correlation is consistent with the interpretation that stronger gamma entrainment was associated with larger stimulation-evoked network reconfiguration. By contrast, resting-state relative power did not differ between PAD-LCR and PAD-HCR across the delta, theta, alpha, beta, low-gamma, and high-gamma bands (all *p* > 0.17), indicating that the subgroup difference was not explained by baseline spectral differences.

The PAD-specific cognitive reserve pattern was similar across several additional analytic approaches. When graph metrics were recomputed at 50%, 70%, and 90% network density thresholds, the PAD-LCR versus PAD-HCR difference remained similar in magnitude, particularly for mean strength (Cohen’s *d* = 0.90, 0.92, and 0.98, respectively). In addition, a non-parametric permutation test with 10,000 random permutations of cognitive reserve group labels yielded significant results for both the within-PAD cognitive reserve effect (*p* = 0.021) and the global diagnosis-by-cognitive reserve interaction in stimulation-related change scores across diagnostic groups (*p* = 0.028).

To confirm that the symmetrization step did not drive the main findings, we recomputed directed-network metrics from the raw asymmetric 32 Hz sGC matrices. Within the PAD group, the LCR subgroup again showed a greater stimulation-related increase in total directed strength than the HCR subgroup (Δ = 546.2 vs. 205.8, *p* = 0.043, *d* = 1.021), closely matching the effect observed for symmetrized mean strength. By contrast, stimulation-related change in network asymmetry did not differ between the PAD cognitive reserve subgroups (*p* = 0.847), suggesting that the cognitive reserve-dependent effect primarily reflected the magnitude of connectivity increase rather than a differential reorganization of flow direction.

Additional sensitivity analyses are reported in **Supplementary Tables S3, S5, and S6**. Leave-one-out-analyses showed that the three-way interaction remained significant for outreach after exclusion of any single PAD-LCR participant but was attenuated for mean strength and weighted clustering coefficient upon removal of one influential participant (**Supplementary Table S3**). When the 12 amyloid-positive cognitively normal participants were included as a fourth diagnostic group, the three-way interaction remained significant for mean strength, weighted clustering coefficient, and outreach (**Supplementary Table S5**). When cognitive reserve was modelled as a continuous variable, the three-way interaction did not reach statistical significance (all *p* > 0.19; **Supplementary Table S6**). Taken together, these analyses suggest that the main pattern was directionally consistent across some alternative specifications but remained sensitive to influential observations and to the way cognitive reserve was modelled. Future studies with larger samples and validated continuous reserve measures are needed to clarify the functional form and robustness of this relationship.

We then conducted exploratory analyses to characterize stimulation-related network response heterogeneity within the PAD group. Although the three-way interaction was not significant for the average shortest path length at the group level, changes in path length can still capture meaningful individual-level variation in functional integration when examined jointly with network strengthening. Accordingly, we evaluated participant-level joint changes in network strength and path-length reduction to describe response patterns that may be obscured in omnibus interaction tests.

**Fig. 2A** plots individual stimulation-related changes in mean strength and average shortest path length (ΔMS and ΔPL), providing a participant-level view of how network strengthening and path-length reduction co-occurred. HCR participants tended to show larger reductions in path length despite relatively small increases in mean strength, whereas LCR participants more often exhibited stronger increases in mean strength with comparatively limited path length reduction. In line with this group difference, the ΔPL/ΔMS ratio differed between CR groups (Welch’s *t*(22.62) = 2.13, *p* = 0.045, 95% CI [0.24, 18.22]). In addition, the outreach/mean strength ratio during stimulation was higher in the LCR group than in the HCR group (*t*(20.84) = 2.24, *p* = 0.036, 95% CI [0.01, 0.07]), suggesting relatively greater outreach in relation to overall strengthening under stimulation.

**Figure 2.**
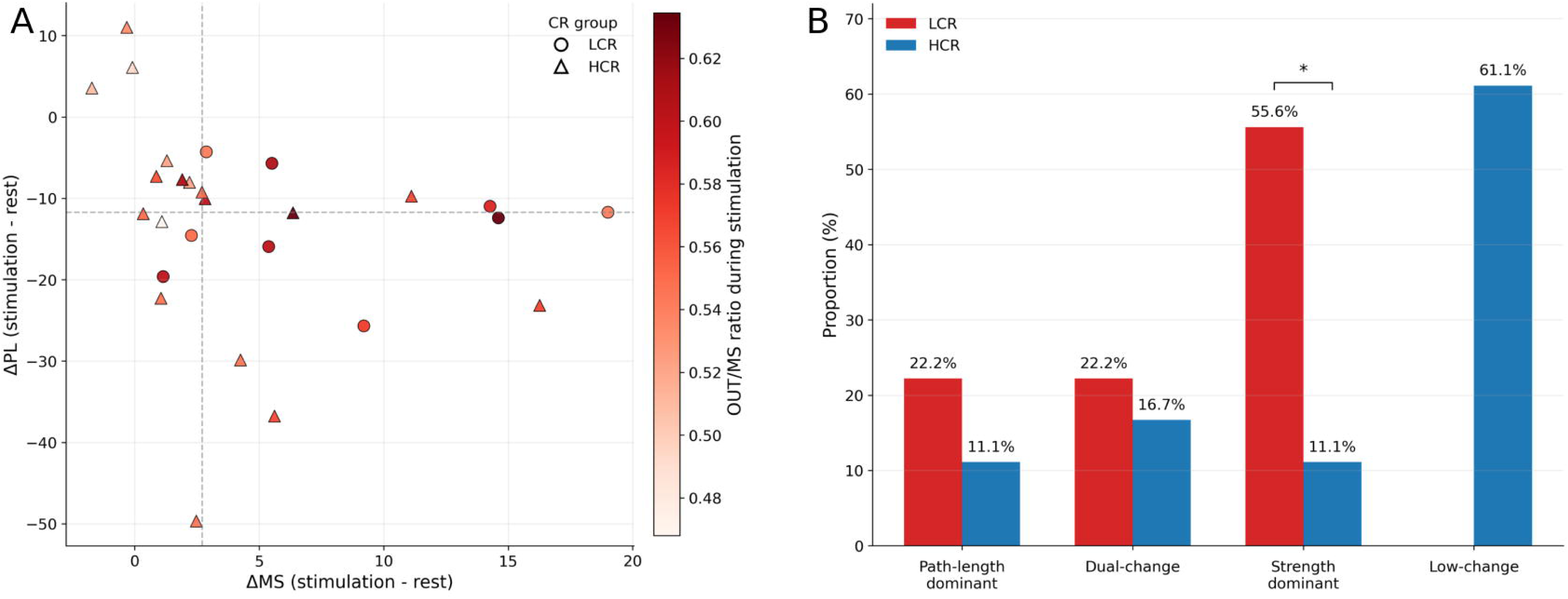
Exploratory visualization of stimulation-related network changes by cognitive reserve in PAD. (A) Individual changes in mean strength (MS) and average shortest path length (PL) are shown by CR group, with ΔMS defined as stimulation – rest and ΔPL defined as rest – stimulation. Marker shape indicates CR group, and marker color represents the Outreach/MS (OUT/MS) ratio during stimulation. Dashed lines denote the median splits of ΔMS and ΔPL used for response-pattern classification. (B) Proportion of participants classified into four exploratory response patterns based on the median splits of ΔMS and ΔPL (path-length-dominant, dual-change, strength-dominant, and low-change). The distribution of response patterns differed between CR groups (Fisher’s exact test, p = 0.026). Group differences in the ΔPL/ΔMS and OUT/MS ratios were also significant (Welch’s t tests, p = 0.045 and p = 0.036, respectively), but these exploratory analyses were not corrected for multiplicity and should be interpreted cautiously. The quadrants in panel B were defined by two axes: ΔMS > 0 (stimulation > rest for mean strength) and ΔPL > 0 (rest > stimulation for average shortest path length, i.e., path-length reduction). The four response patterns were classified as: path-length-dominant (ΔMS ≤ 0 and ΔPL > 0), dual-change (ΔMS > 0 and ΔPL > 0), strength-dominant (ΔMS > 0 and ΔPL ≤ 0), and low-change (ΔMS ≤ 0 and ΔPL ≤ 0).

A complementary exploratory quadrant-based classification of individual responses further highlighted this divergence (**Fig. 2B**). LCR participants were more likely to display a strength-dominant pattern with relatively limited path-length reduction (55.6% vs 11.1%), whereas HCR participants more frequently showed a low-change pattern (61.1% vs 0%). When the classification was collapsed into a binary indicator of the strength-dominant pattern, response profile was significantly associated with cognitive reserve level within the PAD group (*p* = 0.026). These exploratory results should be regarded as hypothesis-generating. Rather than establishing a stable subgroup effect, they suggest the possibility that cognitive reserve may be related not only to the magnitude of stimulation response in PAD, but also to the pattern of network reconfiguration.

## Discussion

In the present study, we examined whether CR was associated with differences in brain network response to 32 Hz visual gamma stimulation across three diagnostic groups. Significant condition-by-diagnosis-by-CR interactions for mean strength, weighted clustering coefficient, and outreach were driven primarily by the PAD group, in which LCR participants showed larger rest-to-stimulation network increases than HCR participants. Several supportive analyses were directionally consistent with this PAD-specific pattern, including higher 32 Hz entrainment in the LCR subgroup, positive correlations between entrainment strength and network change at the whole-sample level, similar results across density thresholds, non-parametric permutation testing, and a directed reanalysis of the raw asymmetric sGC matrices. Nevertheless, this subgroup pattern should be considered preliminary given the very small PAD-LCR subgroup (*n* = 9), the sensitivity of some findings to removal of a single participant, and the absence of a significant three-way interaction when CR was modelled continuously. Taken together, these findings suggest the possibility that cognitive reserve may be associated with heterogeneity in stimulation-related network reconfiguration, particularly in PAD.

The PAD-specific concentration of the CR-dependent stimulation effect may reflect the intermediate neurobiological position of this group between CN and AD. PAD represents a transitional stage in which AD-related pathology is already sufficient to perturb large-scale network organization^38, 39^, yet compensatory network mechanisms may still be sufficiently preserved to support flexible reorganization in response to external stimulation^40^. Notably, resting-state graph metrics did not differ significantly between CR subgroups within any diagnostic group (**Supplementary Table S1**), suggesting that the reserve-related divergence emerged specifically under stimulation rather than reflecting pre-existing differences in baseline network architecture. This is consistent with the hypothesis that cognitive reserve may manifest as a differential capacity for dynamic network reconfiguration rather than as static topological differences^27^. By contrast, amyloid-negative controls may have too little pathological burden for reserve-related differences to manifest as divergent stimulation responses^41–43^, whereas in established AD, advanced network disconnection may constrain the capacity for large-scale reconfiguration^16, 44^. This interpretation is broadly consistent with cognitive reserve theory, which proposes that reserve-related advantages become most apparent when pathology is substantial enough to challenge the system but not yet so advanced as to preclude adaptive recruitment^27^. The prodromal stage may therefore represent a functionally sensitive window in which reserve-related variability in neural adaptability becomes more detectable under stimulation.

The amplified response in PAD-LCR should not be interpreted as a more efficient or beneficial form of network adaptation. Instead, it may reflect a less stable or less efficient mode of reconfiguration, characterized by broad strengthening of network coupling (mean strength), greater local clustering (weighted clustering coefficient), and wider spatial distribution of network engagement (outreach)^21, 45^. By contrast, average shortest path length did not show a significant three-way interaction, indicating that stimulation-related changes in global integration were less clearly differentiated across groups^46^. Exploratory analyses suggested that PAD-HCR participants tended to show greater path-length reduction relative to strength increase, whereas PAD-LCR participants more often showed pronounced strengthening without commensurate path-length reduction. One possible interpretation is that lower reserve may be associated with greater susceptibility to perturbation, leading to an amplified but potentially less efficient response, whereas the HCR pattern may reflect more selective routing reorganization consistent with the neural efficiency hypothesis. However, these interpretations remain speculative given the exploratory analyses and small subgroup sizes.

Across the full sample, stronger entrainment was accompanied by larger stimulation-related network change, suggesting a general link between gamma entrainment and network reorganization, although this association did not reach significance within PAD alone.

Prior studies have demonstrated the feasibility of sensory gamma stimulation in AD but reported considerable interindividual variability^47–49^, and our previous work identified lower gamma frequencies around 32-34 Hz as more effective for entraining and propagating gamma rhythm in older adults^35–37^. The present findings extend this literature by suggesting that heterogeneity in stimulation responses may be partly structured by reserve-related differences in network adaptability, especially in PAD.

Several limitations should be considered when interpreting the present findings. Importantly, the PAD-LCR subgroup comprised only nine participants, which is the primary constraint on the interpretation of the present findings. Although the adjusted effect sizes were larger (d_adjusted = 3.55-3.69; 95% CI [2.57, 6.65]), these estimates were derived from covariate-adjusted residual variance in a small subgroup and should be interpreted with caution. Leave-one-out analyses showed that the three-way interaction was attenuated upon removal of one influential participant (**Supplementary Table S3**), further underscoring the fragility of the subgroup effect. Moreover, the absence of a significant interaction when CR was modelled continuously (**Supplementary Table S6**) suggests that the present subgroup findings should be interpreted cautiously and may depend in part on dichotomization. An exploratory model including the quadratic term of the CR score did not yield a significant three-way interaction (**Supplementary Table S9**), providing no evidence of a non-linear effect. Replication in a larger PAD sample with broader and more continuous reserve measures is therefore essential.

Second, CR was operationalized using a median split of a composite score based on education and occupation attainment, which may not capture the full breadth of the construct, including contributions from cognitively stimulating leisure activities, social engagement, and other lifestyle factors.

Third, several methodological factors warrant consideration. All participants received stimulation at a fixed frequency of 32 Hz rather than an individually optimized frequency, which may have introduced variability in the degree of entrainment achieved. The proportional threshold of 90% network density used in the primary analyses retains a larger number of connections, and although the main pattern was consistent across lower density thresholds, the possibility that weaker connections influenced the absolute values of network metrics cannot be excluded. Additionally, sensor-level EEG connectivity measures may be influenced by the spread of electrical signals across the scalp, and the present findings should be confirmed using source-level analyses. Exploratory analyses were not corrected for multiple comparisons and should be regarded as hypothesis-generating.

In conclusion, the present study raises the preliminary possibility that CR may be related to stimulation-induced network reconfiguration in AD-related cognitive impairment, with the clearest signal observed in PAD. However, the subgroup effect was driven by a small low-reserve PAD subgroup, was sensitive to removal of an influential participant, and was not significant when CR was modelled continuously. Accordingly, these findings should be interpreted cautiously and viewed as hypothesis-generating until replicated in larger independent samples. More broadly, the present results suggest that individual differences in reserve may be relevant when interpreting neural responses to sensory gamma stimulation and may merit explicit consideration in future studies.

## Declarations

### Ethics approval and consent to participate

This study was approved by the Institutional Review Board of Seoul National University Bundang Hospital (IRB No. B-2304-823-001) and was conducted in accordance with the Declaration of Helsinki. All participants provided written informed consent prior to enrolment.

### Consent for publication

Not applicable

### Availability of data and materials

The data that support the findings of this study are available from the corresponding author upon reasonable request. The data are not publicly available due to restrictions imposed by the Institutional Review Board of Seoul National University Bundang Hospital to protect participant privacy.

### Authors’ contributions

M.H. performed analysis and drafted the manuscript. Y.S.P. designed experiments, performed the experiments, analysis, and drafted the manuscript. E.Y. and Y.K. contributed to data acquisition. H.C. and S.Y. designed and developed the visual flicker stimulation device. K.W.K. and J.W.H. designed experiments, drafted, and finalized the manuscript. All authors reviewed the manuscript.

## Data Availability

All data produced in the present study are available upon reasonable request to the authors

## Acknowledgements

Not applicable

