## Supplementary material for "Cognitive reserve and gamma entrainment-related network changes in prodromal Alzheimer’s disease"

**Supplementary Appendix 1. OLED-based FLS Device System**

***Device Design and Configuration***

The FLS device was designed as eyewear containing organic light-emitting diode (OLED) panels positioned 2 cm from each pupil. The glasses-like frame was fabricated using a 3D printer (SLA-type with ABS-like resin; HY3D Co., Ltd., Ansan, Korea). Each eye received light from two OLED panels (A88MA2B; Konica Minolta Inc., Tokyo, Japan) with individual active areas of 43.4 mm × 15.9 mm, arranged along the short axis to create a combined active area of 43.4 mm × 31.8 mm per eye. Due to bus electrode constraints, a 7 mm gap existed between the panels. However, geometric simulation using LightTools™ software confirmed uniform light distribution across the pupil despite this gap (**Supplementary Figure S1**).

***Optical Characteristics***

The OLED panels were characterized using a spectroradiometer (CS2000, Konica Minolta Inc.) and source measure unit (Keithley 2400, Tektronix Inc.). To achieve the target illuminance of 1385 lm/m² at a 2 mm pupil diameter and 2 cm viewing distance, we set the OLED luminance to 1120 cd/m². The emission spectrum ranged from 400-700 nm with a primary peak at 446 nm and secondary peaks at 437 nm and 562 nm, resulting in a color temperature of 5700 K similar to daylight (**Supplementary Figure S2**).

***Thermal Management and Safety Features***

Thermal imaging (U5856A; Keysight Technologies Inc.) showed that panel temperature stabilized at 37°C after 20 minutes of continuous operation, remaining stable for extended periods up to 90 minutes (**Supplementary Fig. S3**). To ensure safety and reliability, the device incorporated several protective features. A battery backup system prevented unintended shutdowns during use, while an automatic power-off relay timer ensured complete shutdown after the designated period.

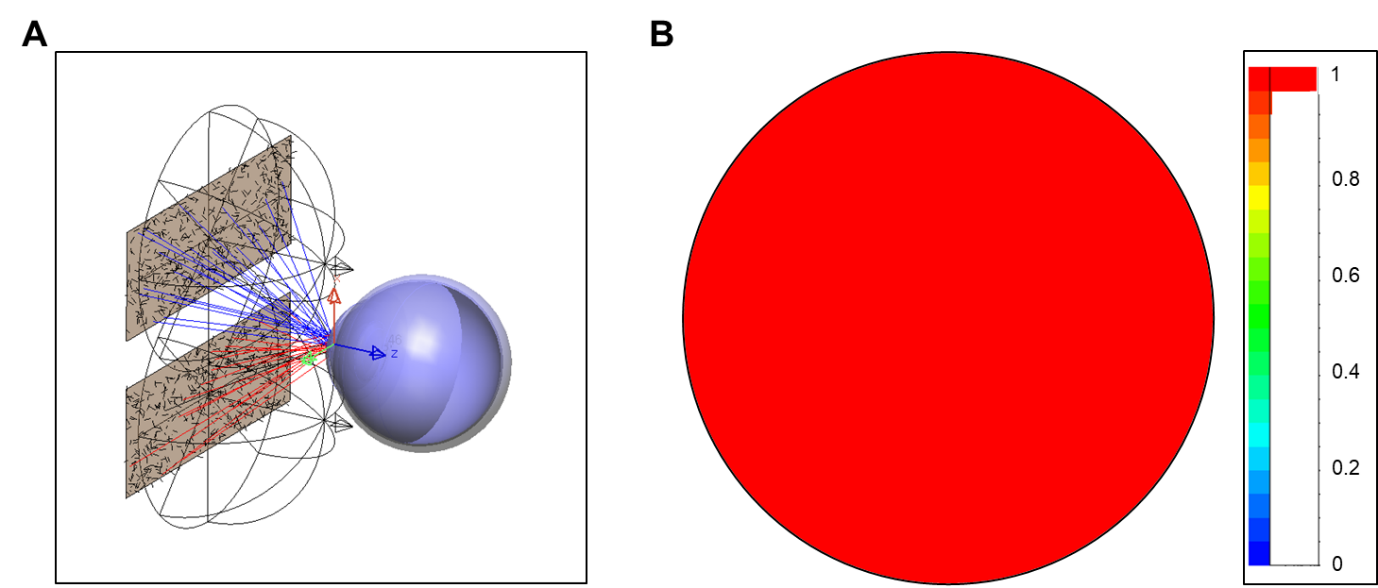

**Supplementary Figure S1**. Light distribution delivered to the pupil by two panels based on geometric optics simulation. (A) A schematic diagram of the eye model (LightTools^TM^) and OLED panels separated by 2 cm. (B) A result of the illuminance distribution at the pupil

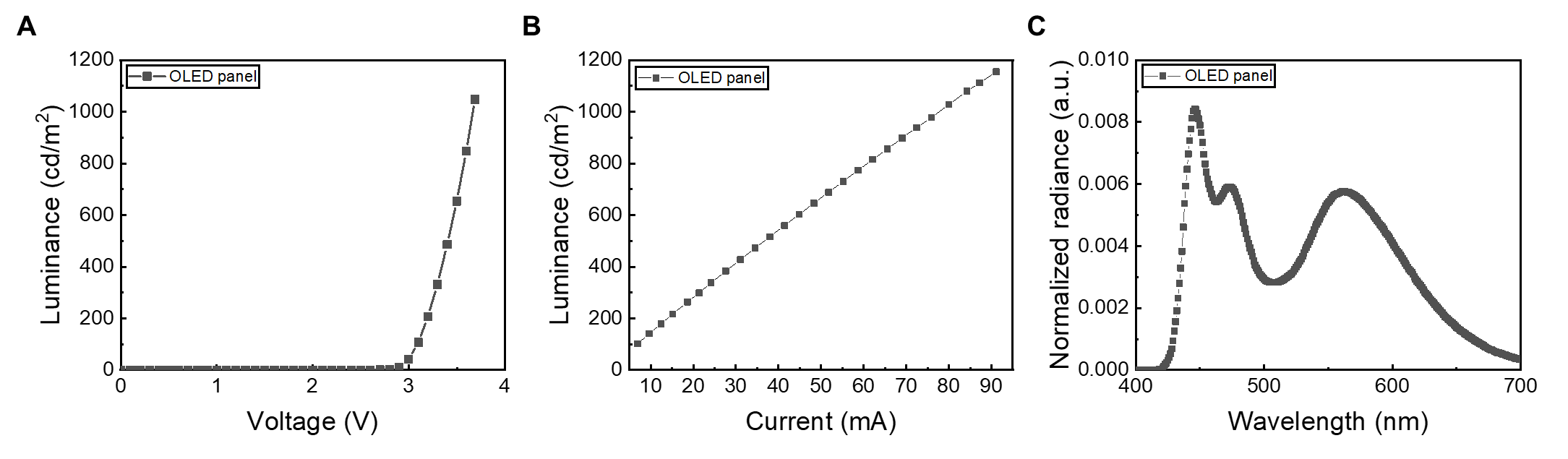

**Supplementary Figure S2**. Electrical and optical characteristics of a single OLED panel: Relationships between luminance and (A) Voltage, (B) Current. (C) Normalized spectrum of white OLED panels with 5700K.

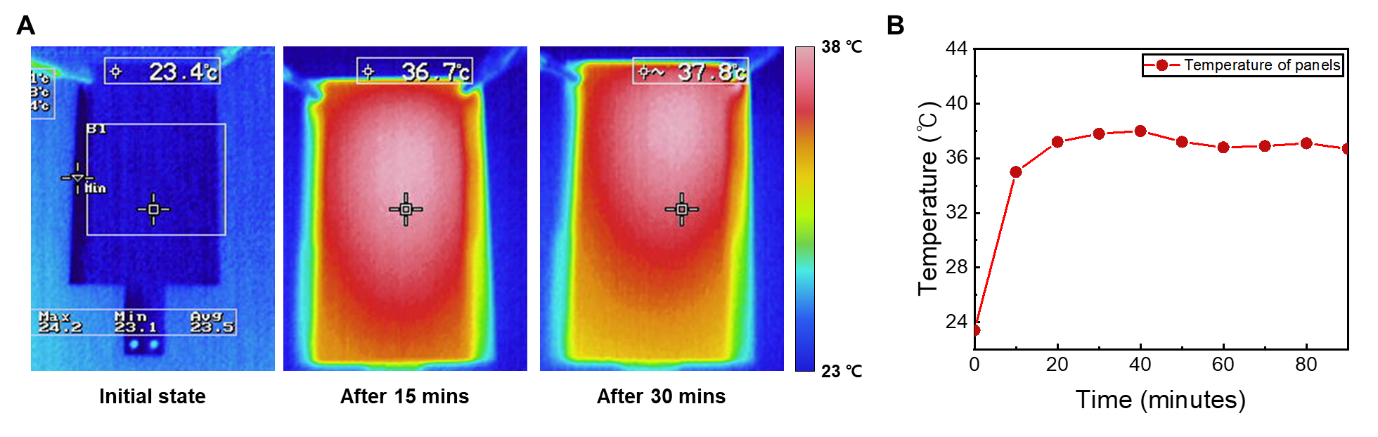

**Supplementary Figure S3**. Temperature changes of the OLED panel during continuous operation at 1120 cd/m²: (A) Thermal camera images over driving time. (B) Temperature variation graph during continuous operation for 90 minutes (3 times of actual experiment duration).

**Supplementary Appendix 2. Unadjusted within-subject effect sizes in PAD subgroups**

For interpretive reference, unadjusted within-subject effect sizes were additionally calculated from the raw observed data within each PAD subgroup using paired-samples t-tests without covariate adjustment. Cohen’s *d* was calculated as *d* = *t*/√*n*. For consistency of interpretation across metrics, mean strength, weighted clustering coefficient, and outreach were defined in the stimulation-minus-rest direction.

Consistent with the adjusted analyses, the unadjusted within-subject effect sizes were larger in the LCR subgroup than in the HCR subgroup for all three metrics (mean strength, 1.30 vs 0.74; weighted clustering coefficient, 1.23 vs 0.70; outreach 1.33 vs 0.74).

**Supplementary Table S1**. **Effects of diagnosis and cognitive reserve on the graph metrics of resting functional connectivity**

|  | MS | | |  | Cw | | |  | PL | | |  |  | Outreach | | |
| --- | --- | --- | --- | --- | --- | --- | --- | --- | --- | --- | --- | --- | --- | --- | --- | --- |
|  | *F* | *p* | *ηp²* |  | *F* | *p* | *ηp²* |  | *F* | *p* | *ηp²* |  |  | *F* | *p* | *ηp²* |
| Diagnosis | 0.717 | 0.491 | 0.016 |  | 0.513 | 0.600 | 0.012 |  | 2.595 | 0.080 | 0.057 |  |  | 0.844 | 0.433 | 0.019 |
| Cognitive reserve | 0.003 | 0.954 | 0.000 |  | 0.026 | 0.873 | 0.000 |  | 0.005 | 0.946 | 0.000 |  |  | 0.035 | 0.853 | 0.000 |
| Cognitive reserve*Diagnosis | 0.215 | 0.807 | 0.005 |  | 0.161 | 0.852 | 0.004 |  | 1.346 | 0.266 | 0.030 |  |  | 0.160 | 0.853 | 0.004 |

**^*^**Analysis of variance examined the main effects of diagnosis (amyloid-negative cognitively normal controls versus prodromal Alzheimer’s disease versus amyloid-positive dementia due to Alzheimer’s disease), cognitive reserve (low versus high), and their interactions on each graph metric.

**Supplementary Table S2**. **Effects of condition, cognitive reserve, and their interaction on the graph metrics of functional connectivity within the PAD**

|  | MS | | |  | Cw | | |  | PL | | |  |  | Outreach | | |
| --- | --- | --- | --- | --- | --- | --- | --- | --- | --- | --- | --- | --- | --- | --- | --- | --- |
|  | *F* | *p* | *ηp²* |  | *F* | *p* | *ηp²* |  | *F* | *p* | *ηp²* |  |  | *F* | *p* | *ηp²* |
| Condition | 21.769 | **<0.001** | 0.497 |  | 22.704 | **<0.001** | 0.508 |  | 0.188 | 0.669 | 0.008 |  |  | 18.568 | **<0.001** | 0.458 |
| Cognitive reserve | 14.453 | **0.001** | 0.396 |  | 14.762 | **0.001** | 0.402 |  | 3.198 | 0.087 | 0.127 |  |  | 14.243 | **0.001** | 0.393 |
| Condition*Cognitive reserve | 15.565 | **0.001** | 0.414 |  | 15.561 | **0.001** | 0.414 |  | 2.773 | 0.110 | 0.112 |  |  | 13.864 | **0.001** | 0.387 |

**Note:** Follow-up analysis of variance examined the main effects of condition (RS; rest versus stimulation) and cognitive reserve group (CR) and their interaction on each graph metric within prodromal Alzheimer’s disease (PAD).

**Supplementary Table S3. Leave-one-out sensitivity analysis of condition-by-diagnosis-by-cognitive reserve interactions after sequential exclusion of PAD-LCR participants.**

|  | Mean Strength | |  | Weighted Clustering Coefficient | |  | Outreach | |
| --- | --- | --- | --- | --- | --- | --- | --- | --- |
| Excluded Subject | *F* | *p* |  | *F* | *p* |  | *F* | *p* |
| None | 4.624 | **0.012** |  | 4.183 | **0.018** |  | 4.828 | **0.010** |
| S009 | 4.869 | **0.010** |  | 4.495 | **0.014** |  | 5.045 | **0.009** |
| S039 | 2.752 | 0.070 |  | 2.173 | 0.120 |  | 3.127 | **0.049** |
| S041 | 5.569 | **0.005** |  | 4.988 | **0.009** |  | 5.893 | **0.004** |
| S043 | 4.804 | **0.011** |  | 4.268 | **0.017** |  | 4.942 | **0.009** |
| S053 | 4.178 | **0.019** |  | 3.721 | **0.028** |  | 4.396 | **0.015** |
| S099 | 3.361 | **0.039** |  | 3.304 | **0.042** |  | 3.322 | **0.041** |
| S100 | 5.960 | **0.004** |  | 5.402 | **0.006** |  | 6.184 | **0.003** |
| S106 | 3.404 | **0.038** |  | 3.208 | **0.045** |  | 3.509 | **0.034** |
| S118 | 5.864 | **0.004** |  | 5.272 | **0.007** |  | 6.225 | **0.003** |

**Supplementary Note:** Participant S039, whose exclusion attenuated the three-way interaction, was a 62-year-old female with a CR score of 6, MMSE of 15, and homozygous APOE ε4/ε4 carrier status. This participant’s 32 Hz SSVEP SNR was 6.099 (PAD-LCR mean: 2.729) and stimulation-related ΔMS was 19.006 (PAD-LCR mean: 8.252). Although these values fell within the observed range of the LCR subgroup, they represented the highest values, indicating that S039 was influential within this subgroup rather than an obvious outlier. Accordingly, the leave-one-out findings should be interpreted as evidence of subgroup fragility rather than as the detection of an anomalous case. **Supplementary Table S4.** **Effects of condition, diagnosis, and cognitive reserve on the graph metrics of functional connectivity at 34 Hz**

|  | MS | | |  | Cw | | |  | PL | | |  |  | Outreach | | |
| --- | --- | --- | --- | --- | --- | --- | --- | --- | --- | --- | --- | --- | --- | --- | --- | --- |
|  | *F* | *p* | *ηp²* |  | *F* | *p* | *ηp²* |  | *F* | *p* | *ηp²* |  |  | *F* | *p* | *ηp²* |
| Condition | 3.291 | 0.073 | 0.037 |  | 3.975 | **0.049** | 0.044 |  | 0.170 | 0.681 | 0.002 |  |  | 2.703 | 0.104 | 0.030 |
| Diagnosis | 6.398 | **0.003** | 0.130 |  | 5.265 | **0.007** | 0.109 |  | 4.502 | **0.014** | 0.095 |  |  | 7.244 | **0.001** | 0.144 |
| Cognitive reserve | 2.142 | 0.147 | 0.024 |  | 2.082 | 0.153 | 0.024 |  | 0.310 | 0.579 | 0.004 |  |  | 2.277 | 0.135 | 0.026 |
| Condition*Diagnosis | 4.927 | **0.009** | 0.103 |  | 4.406 | **0.015** | 0.093 |  | 0.901 | 0.410 | 0.021 |  |  | 5.554 | **0.005** | 0.114 |
| Condition*Cognitive reserve | 2.505 | 0.117 | 0.028 |  | 2.370 | 0.127 | 0.027 |  | 1.319 | 0.254 | 0.015 |  |  | 2.376 | 0.127 | 0.027 |
| Diagnosis*Cognitive reserve | 2.528 | 0.086 | 0.056 |  | 1.877 | 0.159 | 0.042 |  | 1.156 | 0.320 | 0.026 |  |  | 2.884 | 0.051 | 0.063 |
| Condition*Diagnosis*Cognitive reserve | 3.039 | 0.053 | 0.066 |  | 2.609 | 0.079 | 0.057 |  | 0.980 | 0.380 | 0.022 |  |  | 3.197 | **0.046** | 0.069 |

**Abbreviations:** MS, mean strength; Cw, weighted clustering coefficient; PL, average shortest path length

**^*^**Analysis of variance examined the main effects of condition (rest versus stimulation), diagnosis (amyloid-negative cognitively normal controls versus prodromal Alzheimer’s disease versus amyloid-positive dementia due to Alzheimer’s disease), and cognitive reserve (low versus high), and their interactions on each graph metric.

**Supplementary Table S5. Sensitivity analysis including amyloid-positive cognitively normal participants as a separate diagnostic group for the effects of condition, diagnosis, and cognitive reserve on the graph metrics of functional connectivity at 32 Hz**

|  | MS | | |  | Cw | | |  | PL | | |  |  | Outreach | | |
| --- | --- | --- | --- | --- | --- | --- | --- | --- | --- | --- | --- | --- | --- | --- | --- | --- |
|  | *F* | *p* | *ηp²* |  | *F* | *p* | *ηp²* |  | *F* | *p* | *ηp²* |  |  | *F* | *p* | *ηp²* |
| Condition | 2.534 | 0.115 | 0.026 |  | 3.112 | 0.081 | 0.031 |  | 0.408 | 0.524 | 0.004 |  |  | 2.042 | 0.156 | 0.021 |
| Diagnosis | 4.190 | **0.008** | 0.116 |  | 3.544 | **0.017** | 0.100 |  | 2.124 | 0.102 | 0.062 |  |  | 4.730 | **0.004** | 0.129 |
| Cognitive reserve | 0.496 | 0.483 | 0.005 |  | 0.619 | 0.433 | 0.006 |  | 0.384 | 0.537 | 0.004 |  |  | 0.446 | 0.506 | 0.005 |
| Condition*Diagnosis | 3.666 | **0.015** | 0.103 |  | 3.384 | **0.021** | 0.096 |  | 0.594 | 0.621 | 0.018 |  |  | 4.067 | **0.009** | 0.113 |
| Condition*Cognitive reserve | 3.558 | 0.062 | 0.036 |  | 3.480 | 0.065 | 0.035 |  | 4.549 | **0.035** | 0.045 |  |  | 3.221 | 0.076 | 0.032 |
| Diagnosis*Cognitive reserve | 3.259 | **0.025** | 0.092 |  | 2.747 | **0.047** | 0.079 |  | 2.311 | 0.081 | 0.067 |  |  | 3.753 | **0.013** | 0.105 |
| Condition*Diagnosis*Cognitive reserve | 3.231 | **0.026** | 0.092 |  | 2.931 | **0.037** | 0.084 |  | 1.993 | 0.120 | 0.059 |  |  | 3.368 | **0.022** | 0.095 |

**Abbreviations:** MS, mean strength; Cw, weighted clustering coefficient; PL, average shortest path length

**^*^**Analysis of variance examined the main effects of condition (rest versus stimulation), diagnosis (amyloid-negative cognitively normal controls versus amyloid-positive cognitively normal versus prodromal Alzheimer’s disease versus amyloid-positive dementia due to Alzheimer’s disease), and cognitive reserve (low versus high), and their interactions on each graph metric.

**Supplementary Table S6. Effects of condition, diagnosis, and continuous cognitive reserve modeled as a continuous variable on functional connectivity graph metrics at 32 Hz**

|  | MS | | |  | Cw | | |  | PL | | |  |  | Outreach | | |
| --- | --- | --- | --- | --- | --- | --- | --- | --- | --- | --- | --- | --- | --- | --- | --- | --- |
|  | *F* | *p* | *ηp²* |  | *F* | *p* | *ηp²* |  | *F* | *p* | *ηp²* |  |  | *F* | *p* | *ηp²* |
| Condition | 3.552 | 0.063 | 0.040 |  | 4.391 | 0.039 | 0.049 |  | 0.829 | 0.365 | 0.010 |  |  | 2.871 | 0.094 | 0.032 |
| Diagnosis | 3.120 | **0.049** | 0.068 |  | 2.625 | 0.078 | 0.058 |  | 5.402 | **0.006** | 0.112 |  |  | 3.438 | **0.037** | 0.074 |
| Cognitive reserve score | 2.143 | 0.147 | 0.024 |  | 2.166 | 0.145 | 0.025 |  | 2.189 | 0.143 | 0.025 |  |  | 1.949 | 0.166 | 0.022 |
| Condition*Diagnosis | 2.487 | 0.089 | 0.055 |  | 2.172 | 0.120 | 0.048 |  | 1.403 | 0.251 | 0.032 |  |  | 2.666 | 0.075 | 0.058 |
| Condition*Cognitive reserve score | 0.914 | 0.342 | 0.011 |  | 1.804 | 0.301 | 0.012 |  | 0.317 | 0.575 | 0.004 |  |  | 0.714 | 0.400 | 0.008 |
| Diagnosis*Cognitive reserve score | 1.899 | 0.156 | 0.042 |  | 1.604 | 0.207 | 0.036 |  | 4.005 | 0.022 | 0.085 |  |  | 2.091 | 0.130 | 0.046 |
| Condition*Diagnosis*Cognitive reserve score | 1.581 | 0.212 | 0.035 |  | 1.342 | 0.267 | 0.030 |  | 1.401 | 0.252 | 0.032 |  |  | 1.685 | 0.191 | 0.038 |

**Abbreviations:** MS, mean strength; Cw, weighted clustering coefficient; PL, average shortest path length

**^*^**Analysis of variance examined the main effects of condition (rest versus stimulation), diagnosis (amyloid-negative cognitively normal controls versus prodromal Alzheimer’s disease versus amyloid-positive dementia due to Alzheimer’s disease), and cognitive reserve score, and their interactions on each graph metric.

**Supplementary Table S7. SSVEP signal-to-noise ratio at 32 Hz according to diagnostic stage and cognitive reserve subgroup**

|  | LCR *n* | LCR Mean (SD) | HCR *n* | HCR Mean (SD) | Statistics |  |  |
| --- | --- | --- | --- | --- | --- | --- | --- |
|  |  |  |  |  | Welch’s *t* | *p* | Cohen’s *d* |
| CN | 6 | 1.987 (0.524) | 20 | 2.397 (1.340) | -1.114 | 0.278 | -0.403 |
| PAD | 9 | 2.729 (1.516) | 18 | 1.230 (1.435) | 2.465 | **0.026** | 1.015 |
| AD | 23 | 1.240 (1.484) | 19 | 1.336 (1.299) | -0.224 | 0.824 | -0.069 |

**Abbreviations:** CN, amyloid-negative cognitively normal controls; PAD, prodromal Alzheimer’s disease defined as amyloid-positive mild cognitive impairment; AD, amyloid-positive dementia due to Alzheimer’s disease; LCR, low cognitive reserve; HCR, high cognitive reserve.

**Note:** Welch’s *t*-tests were conducted within each diagnostic group.

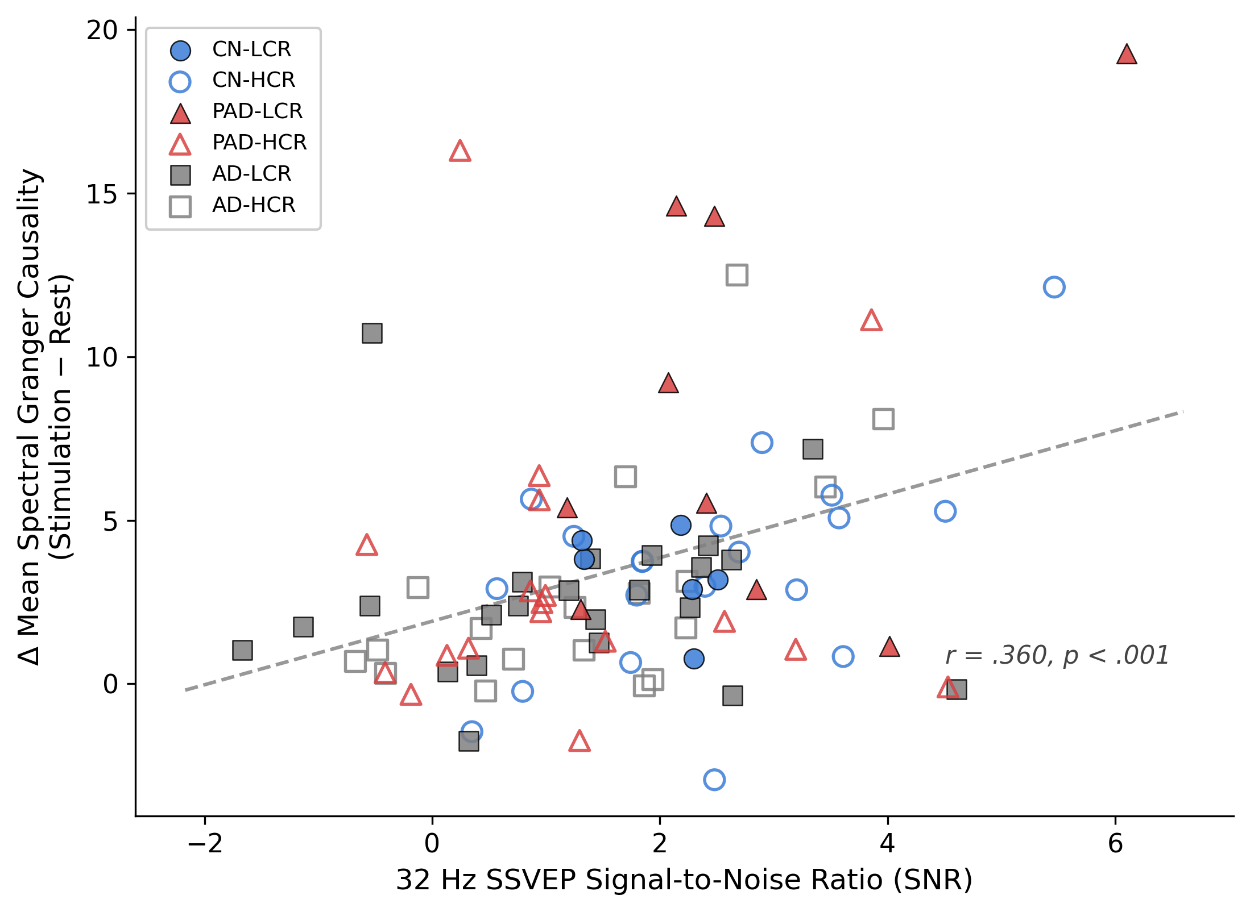

**Supplementary Figure S4. Association between 32 Hz entrainment strength and stimulation-related network change.** Scatter plot of 32 Hz SSVEP SNR versus stimulation-related changes in mean sGC (Δ sGC = stimulation – rest) at 32 Hz. Marker shape indicates diagnostic group, and fill indicates cognitive reserve group. The dashed line represents the overall linear regression across all 95 participants (*r* = 0.360, *p* < 0.001). Per-group Pearson correlations are shown in the main text.

**Abbreviations:** CN, amyloid-negative cognitively normal controls; PAD, prodromal Alzheimer’s disease; AD, amyloid-positive dementia due to Alzheimer’s disease; LCR, low cognitive reserve; HCR, high cognitive reserve.

**Supplementary Table S8. Directed-network reanalysis of stimulation-related changes in 32 Hz sGC metrics according to diagnostic stage and cognitive reserve subgroup.**

|  |  | LCR *n* | LCR Mean (SD) | HCR *n* | HCR Mean (SD) | Statistics |  |  |
| --- | --- | --- | --- | --- | --- | --- | --- | --- |
|  | |  |  |  |  | Welch’s *t* | *p* | Cohen’s *d* |
| Δ Total Directed Strength | |  |  |  |  |  |  |  |
|  | CN | 6 | 209.1 (90.8) | 20 | 221.9 (206.8) | -0.217 | 0.831 | -0.080 |
|  | PAD | 9 | 546.2 (413.0) | 18 | 205.8 (227.3) | 2.304 | 0.043 | 1.021 |
|  | AD | 23 | 180.8 (202.1) | 19 | 170.9 (196.8) | 0.161 | 0.873 | 0.050 |
| Δ Network Asymmetry | |  |  |  |  |  |  |  |
|  | CN | 6 | 0.06 (0.06) | 20 | 0.05 (0.13) | 0.253 | 0.803 | 0.093 |
|  | PAD | 9 | 0.01 (0.18) | 18 | 0.03 (0.11) | -0.198 | 0.847 | -0.086 |
|  | AD | 23 | 0.01 (0.13) | 19 | 0.02 (0.11) | -0.264 | 0.793 | -0.081 |
| Δ Net Flow Variance | |  |  |  |  |  |  |  |
|  | CN | 6 | 28.4 (20.1) | 20 | 28.9 (28.4) | -0.055 | 0.957 | -0.023 |
|  | PAD | 9 | 110.5 (131.7) | 18 | 35.3 (71.6) | 1.598 | 0.140 | 0.709 |
|  | AD | 23 | 26.8 (65.3) | 19 | 25.4 (49.0) | 0.074 | 0.941 | 0.023 |
| Δ Mean Out-Strength | |  |  |  |  |  |  |  |
|  | CN | 6 | 3.32 (1.44) | 20 | 3.52 (3.28) | -0.217 | 0.831 | -0.080 |
|  | PAD | 9 | 8.67 (6.56) | 18 | 3.27 (3.61) | 2.304 | 0.043 | 1.021 |
|  | AD | 23 | 2.87 (3.21) | 19 | 2.71 (3.12) | 0.161 | 0.873 | 0.050 |

**Abbreviations:** CN, amyloid-negative cognitively normal controls; PAD, prodromal Alzheimer’s disease defined as amyloid-positive mild cognitive impairment; AD, amyloid-positive dementia due to Alzheimer’s disease; LCR, low cognitive reserve; HCR, high cognitive reserve.

**Note:** Values represent stimulation minus rest change scores derived from raw asymmetric 32 Hz sGC matrices. Directed metrics included total directed strength, mean out-strength, network asymmetry, and net flow variance. Welch’s *t*-tests were conducted to compare LCR and HCR within each diagnostic group. Positive Δ values indicate larger metric values during stimulation than during rest within the corresponding subgroup. These directed comparisons are presented as supportive sensitivity analyses.

**Supplementary Table S9. Exploratory quadratic cognitive reserve model: effects of condition, diagnosis, and the quadratic term of continuous cognitive reserve on functional connectivity graph metrics at 32 Hz.**

|  | MS | | |  | Cw | | |  | PL | | |  | Outreach | | |
| --- | --- | --- | --- | --- | --- | --- | --- | --- | --- | --- | --- | --- | --- | --- | --- |
|  | *F* | *p* | *ηp²* |  | *F* | *p* | *ηp²* |  | *F* | *p* | *ηp²* |  | *F* | *p* | *ηp²* |
| Condition $\times$ Diagnosis $\times$ CR² | 0.275 | 0.760 | 0.006 |  | 0.357 | 0.701 | 0.008 |  | 0.605 | 0.549 | 0.012 |  | 0.216 | 0.806 | 0.005 |

**Abbreviations:** MS, mean strength; Cw, weighted clustering coefficient; PL, average shortest path length; CR², quadratic term of centered cognitive reserve score.

**Note:** Repeated-measure ANCOVA with condition (rest versus stimulation) as a within-subject factor and diagnosis (CN, PAD, AD) as a between-subject factor, with age, sex, APOE ε4 carrier status, centered CR scores, and the quadratic term of centered CR score (CR²) as covariates. The three-way interaction of condition $\times$ diagnosis $\times$ CR² tests whether the relationship between cognitive reserve and stimulation-related network change varies non-linearly across diagnostic groups. In the current sample, the non-significant three-way interaction provided no evidence for a quadratic reserve effect.

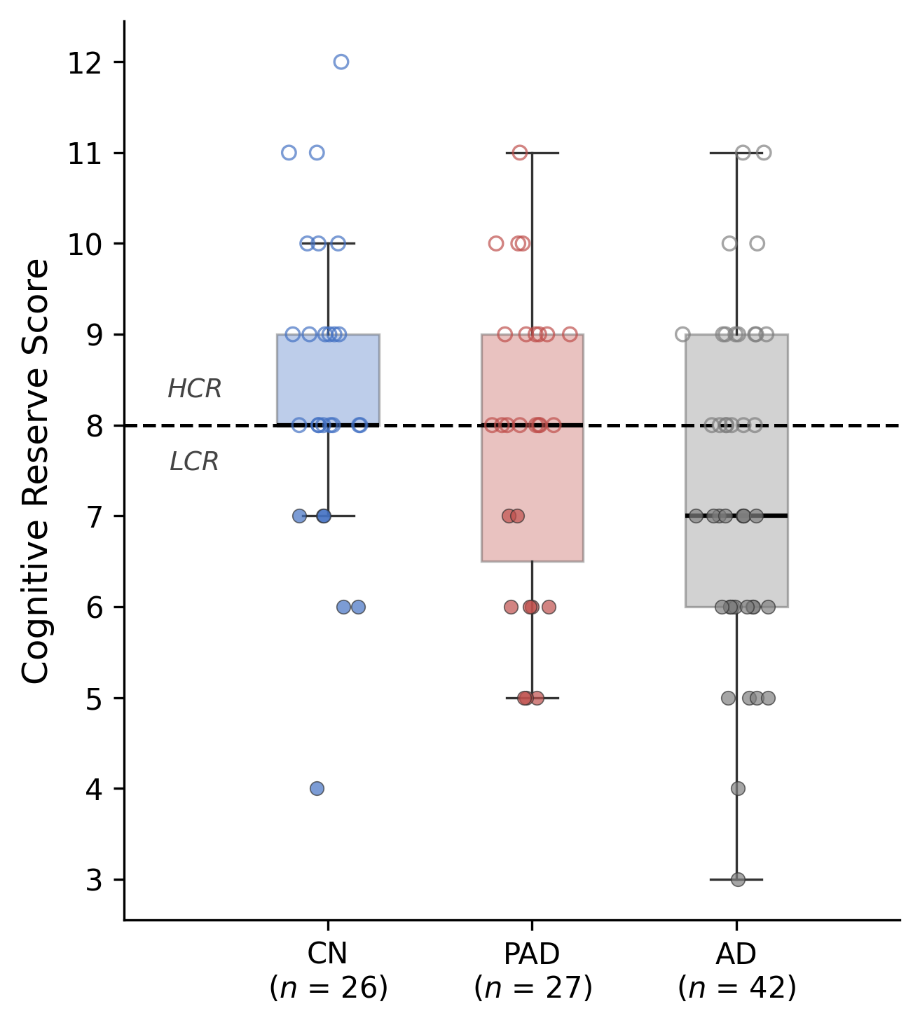

**Supplementary Figure S5. Distribution of cognitive reserve scores across diagnostic groups.** Box-and-strip plots showing cognitive reserve score distributions for the full sample and each diagnostic group. Individual data points are jittered horizontally; filled markers indicate LCR and open markers indicate HCR. The dashed line indicates the median split cutoff (score = 8) used for exploratory subgroup classification in the main analyses.
